# Intention to have the seasonal influenza vaccination during the COVID-19 pandemic among eligible adults in the UK

**DOI:** 10.1101/2020.10.26.20219592

**Authors:** Susan M. Sherman, Julius Sim, Richard Amlôt, Megan Cutts, Hannah Dasch, G James Rubin, Nick Sevdalis, Louise E. Smith

## Abstract

We investigated likelihood of having the seasonal influenza vaccination in 645 participants who were eligible for the vaccination in the UK. 55.8% indicated they were likely to have the vaccination. Previous research suggests that increasing uptake of the influenza vaccination may help contain a COVID-19 outbreak, so steps need to be taken to convert intention into behaviour and to reach the 23.9% who were unlikely to have the vaccination and the 20.3% who were unsure.

---

As the seasonal influenza season in Europe and the US approaches, we need to understand any impact that COVID-19 may have had on intention to have the influenza vaccination this year in order to maximize uptake and help to contain subsequent COVID-19 outbreaks. We report findings from a survey conducted in July 2020, which explored participants’ likelihood of having the seasonal influenza vaccination this winter (2020/21).

The COVID-19 pandemic was declared on March 11^th^, 2020. While the first wave of the pandemic missed most of the influenza season in Europe and the US, a second wave is likely to overlap with the 2020–2021 season^1^. Healthcare systems already come under considerable strain during a typical influenza season, which would be compounded if there is an increase in COVID-19 cases this year. Recent research has modelled the impact of mass influenza vaccination on the spread of COVID-19 should such an overlap occur, and the findings suggest that increasing uptake of the influenza vaccination would facilitate efforts to contain a COVID-19 outbreak^2^. However, increasing, or even maintaining, levels of influenza vaccination may be problematic if reduced uptake patterns seen already in other vaccines also hold for the influenza vaccine. For example, the uptake of the measles-mumps-rubella (MMR) vaccine in England became 19.8% lower in the 3 weeks after full physical distancing measures were introduced in March than in 2019^3^.

The influenza season in the UK runs from December until March each year and the national vaccination programme starts from September. The vaccination is available free through the NHS to children aged two to eleven, adults over 65, pregnant women, healthcare workers and individuals aged 6 months to 65 years who are in clinical at-risk groups (many of which coincide with the COVID-19 at-risk groups). The vaccination is also available privately through primary care and pharmacies. Despite the wide availability of a free vaccine for eligible individuals, uptake varies across the different categories of eligibility; for example, 72.4% of 65+ adults in England were vaccinated in the 2019–2020 season compared to 44.9% of individuals aged 6 months to 65 in clinical at-risk groups^4^.

In order to protect people as we approach the next influenza season, it is helpful to understand the impact that the COVID-19 pandemic may have had on intention to have a seasonal influenza vaccination. To this end, we explored participants’ likelihood of having the seasonal influenza vaccination as part of a larger cross-sectional study investigating attitudes towards a potential COVID-19 vaccination^5^.

A nationally representative sample of 1500 UK adults (quotas set on age, gender and ethnicity) were recruited through Prolific’s online research panel to complete a survey that included sociodemographic questions, clinical questions, questions about COVID-19, and questions about a possible COVID-19 vaccination. We also asked participants if they had been vaccinated for seasonal influenza last winter (yes/no), and how likely they would be to have the seasonal influenza vaccine this winter (eleven point scale, from “extremely unlikely” to “extremely likely”). Full details of the wider study, including survey methodology and ethics approval, are reported elsewhere^5^.

645 individuals in our sample were eligible for the influenza vaccine. The distribution of influenza vaccination intention was bi-modal, with the majority of responses clustering at both ends of the scale. We therefore dichotomized this variable as 0–2 = ‘no’ (*n* = 154) and 8–10 = ‘yes’ (*n* = 360) on the 0–10 scale and the 131 indeterminate cases were not analysed further. Of the 514 eligible respondents who expressed a clear intention to have the influenza vaccine or not, 55.8% (*n*=286) were female and 44.2% (*n*=227) were male; mean age was 49.9 (SD=17.5, range from 18 to 87 years); and 87.7% (*n*=448) were white. In addition, 60.6% (*n*=311) reported having the influenza vaccination last winter and 39.4% (*n*=202) reported not having it (denominators for these percentages range from 511 to 514, due to missing values). We subsequently investigated variables associated with intention to receive a seasonal influenza vaccine in 2020/21 using logistic regression analyses, and included predictors informed by previous research:^6,7^ sociodemographic variables; uptake of influenza vaccine last winter; and beliefs about vaccination (value of vaccination in general; not afraid of needles; see Table 1).

**TABLE 1.**
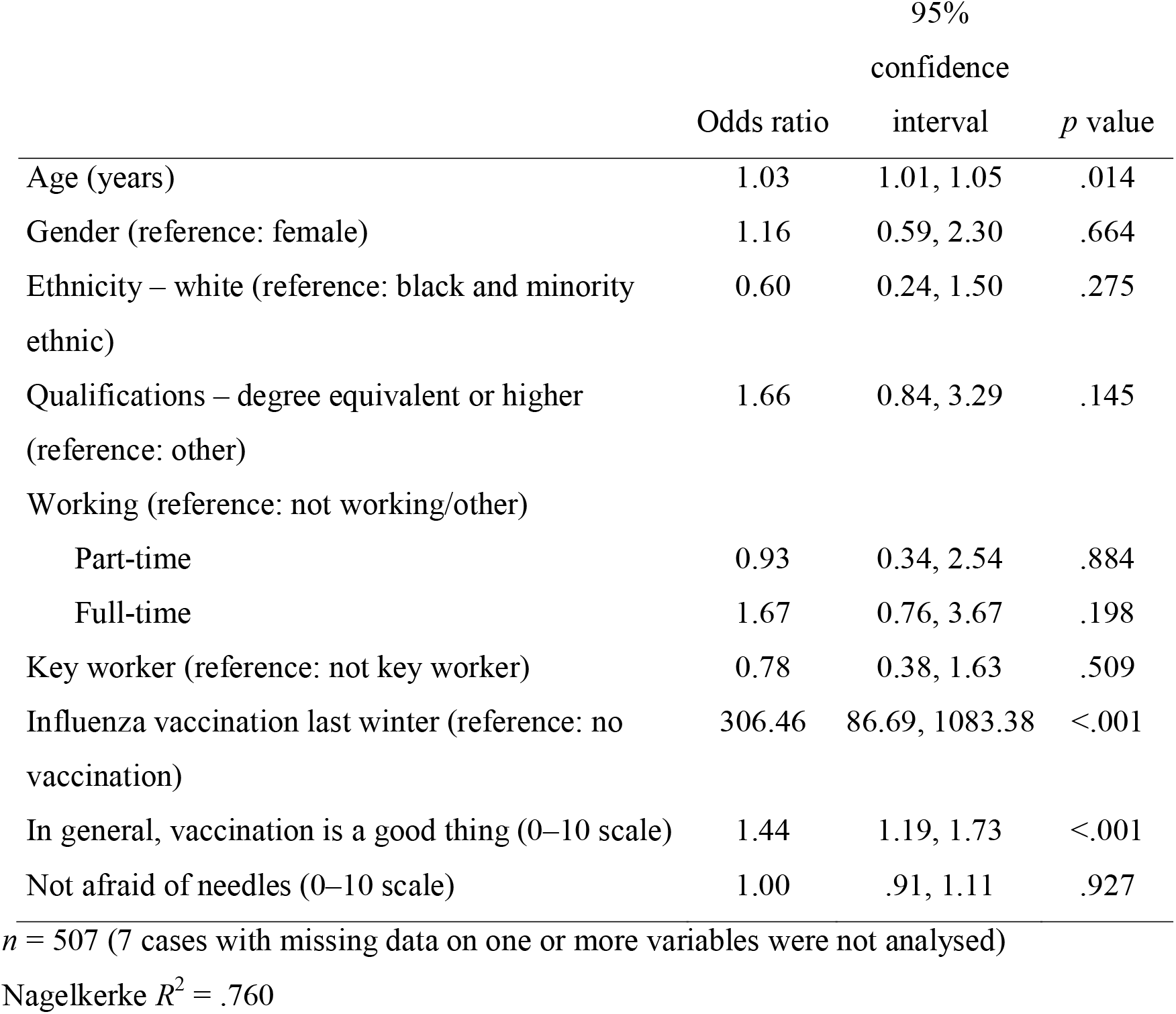
Logistic regression analysis (odds ratios and 95% confidence intervals) of variables associated with intention to receive a seasonal influenza vaccine. Statistical significance was set at *p* ≤.05.

Both age and a positive attitude to vaccination in general were predictors of intention to have the influenza vaccine this year. 64.0% of adults aged 64 or younger who stated a clear intention indicated they would have the vaccination compared to 82.4% of adults aged 65+. However, the strongest predictor by far, as indicated by the large odds ratio, was previous influenza vaccination behaviour.

These findings strongly suggest that individuals who had the influenza vaccine last year are likely to intend to have it again this year, and this is consistent with findings from the H1N1 influenza pandemic^6^. However, there are still key issues to address. Vaccination intention across all those individuals in our sample who were eligible for the vaccine (55.8%) was higher than the reported uptake from last year (32.3%). However, it is likely that actual uptake will be lower than intention as a result of the intention-behaviour gap^8^, making it important that efforts are made to convert positive intentions into uptake. This might be achieved through appropriate messaging and special arrangements for vaccine delivery, particularly for those who might be shielding or at higher risk from COVID-19 and reluctant to attend their GP surgery. Both approaches are also likely to be needed to motivate those individuals who have not previously had the influenza vaccine and those individuals who are eligible for free vaccination but who were among the 44.2% of our sample that indicated they definitely did not intend to be vaccinated (23.9%) or were unsure (20.3%).

The NHS is often overwhelmed during the influenza season, needing, for example, to cancel routine operations. During the first wave of the COVID-19 pandemic in the UK, which occurred outside the influenza season, there was sufficient concern about the NHS’s ability to cope that 10 new ‘Nightingale’ hospitals were built. Increasing uptake of the seasonal influenza vaccine in a timely fashion will relieve pressure on the service. If this is to be successful, strategies to achieve this increase need to be implemented now.

## Data Availability

The link provided is a read-only link. On acceptance this will be updated to a fully accessible link to the data.

https://osf.io/94856/?view_only=c85fd2666a204c67b2c41f0ded105ec2

## Conflict of interest

NS is the director of the London Safety and Training Solutions Ltd, which offers training in patient safety, implementation solutions and human factors to healthcare organisations. The other authors have no conflicts of interest to declare.

## Funding statement

Data collection was funded by a Keele University Faculty of Natural Sciences Research Development award to SS, JS and NS, and a King’s Together Rapid COVID-19 award granted jointly to LS, GJR, RA, NS, SS and JS. LS, RA and GJR are supported by the National Institute for Health Research Health Protection Research Unit (NIHR HPRU) in Emergency Preparedness and Response, a partnership between Public Health England, King’s College London and the University of East Anglia. NS’ research is supported by the National Institute for Health Research (NIHR) Applied Research Collaboration (ARC) South London at King’s College Hospital NHS Foundation Trust. NS is a member of King’s Improvement Science, which offers co-funding to the NIHR ARC South London and comprises a specialist team of improvement scientists and senior researchers based at King’s College London. Its work is funded by King’s Health Partners (Guy’s and St Thomas’ NHS Foundation Trust, King’s College Hospital NHS Foundation Trust, King’s College London and South London and Maudsley NHS Foundation Trust), Guy’s and St Thomas’ Charity and the Maudsley Charity. The views expressed are those of the authors and not necessarily those of the NIHR, the charities, Public Health England or the Department of Health and Social Care.

## Notes

### Author Declarations

Ethical approval for this study was granted by Keele Universitys Research Ethics Committee (reference PS-200129).

## References

1. Maltezou, H. C., Theodoridou, K., & Poland, G. (2020). Influenza immunization and COVID-19. Vaccine, 38(39), 6078–6079.

2. Li, Q., Tang, B., Bragazzi, N. L., Xiao, Y., & Wu, J. (2020). Modeling the impact of mass influenza vaccination and public health interventions on COVID-19 epidemics with limited detection capability. Mathematical Biosciences, 325, 108378.

3. McDonald, H. I., Tessier, E., White, J. M., Woodruff, M., Knowles, C., Bates, C., … & Yarwood, J. (2020). Early impact of the coronavirus disease (COVID-19) pandemic and physical distancing measures on routine childhood vaccinations in England, January to April 2020. Eurosurveillance, 25(19), 2000848.

4. Public Health England (2020). Surveillance of influenza and other respiratory viruses in the UK: Winter 2019 to 2020. London.

5. Sherman, S. M., Smith, L. E., Sim, J., Amlôt, R., Cutts, M., Dasch, H., Rubin, G. R., & Sevdalis, N. (2020). COVID-19 vaccination intention in the UK: Results from the COVID-19 Vaccination Acceptability Study (CoVAccS), a nationally representative cross-sectional survey. medRxiv.

6. Bish, A., Yardley, L., Nicoll, A., & Michie, S. (2011). Factors associated with uptake of vaccination against pandemic influenza: a systematic review. Vaccine, 29(38), 6472–6484.

7. Wheelock A, Miraldo M, Thomson A, Vincent C, Sevdalis N. Evaluating the importance of policy amenable factors in explaining influenza vaccination: a cross-sectional multinational study. BMJ Open 2017;7:e014668

8. Sniehotta, F. F., Scholz, U., & Schwarzer, R. (2005). Bridging the intention–behaviour gap: Planning, self-efficacy, and action control in the adoption and maintenance of physical exercise. Psychology & Health, 20(2), 143–160.

